# Generative AI and Large Language Models in Reducing Medication Related Harm and Adverse Drug Events – A Scoping Review

**DOI:** 10.1101/2024.09.13.24313606

**Authors:** Jasmine Chiat Ling Ong, Chen Michael, Ning Ng, Kabilan Elangovan, Nichole Yue Ting Tan, Liyuan Jin, Qihuang Xie, Daniel Shu Wei Ting, Rosa Rodriguez-Monguio, David W. Bates, Nan Liu

## Abstract

**Background:** Medication-related harm has a significant impact on global healthcare costs and patient outcomes, accounting for deaths in 4.3 per 1000 patients. Generative artificial intelligence (GenAI) has emerged as a promising tool in mitigating risks of medication-related harm. In particular, large language models (LLMs) and well-developed generative adversarial networks (GANs) showing promise for healthcare related tasks. This review aims to explore the scope and effectiveness of generative AI in reducing medication-related harm, identifying existing development and challenges in research.

**Methods:** We searched for peer reviewed articles in PubMed, Web of Science, Embase, and Scopus for literature published from January 2012 to February 2024. We included studies focusing on the development or application of generative AI in mitigating risk for medication-related harm during the entire medication use process. We excluded studies using traditional AI methods only, those unrelated to healthcare settings, or concerning non-prescribed medication uses such as supplements. Extracted variables included study characteristics, AI model specifics and performance, application settings, and any patient outcome evaluated.

**Findings:** A total of 2203 articles were identified, and 14 met the criteria for inclusion into final review. We found that generative AI and large language models were used in a few key applications: drug-drug interaction identification and prediction; clinical decision support and pharmacovigilance. While the performance and utility of these models varied, they generally showed promise in areas like early identification and classification of adverse drug events and support in decision-making for medication management. However, no studies tested these models prospectively, suggesting a need for further investigation into the integration and real-world application of generative AI tools to improve patient safety and healthcare outcomes effectively.

**Interpretation:** Generative AI shows promise in mitigating medication-related harms, but there are gaps in research rigor and ethical considerations. Future research should focus on creation of high-quality, task-specific benchmarking datasets for medication safety and real-world implementation outcomes.

## Introduction

Medication-related harm poses significant health and economic burden globally. The global prevalence of medication related harm was 12%, of which 15% was severe and fatal, causing a mortality rate of up 4.3 per 1000 patients.^1^ In contrast, cardiovascular deaths caused by ischemic heart disease accounted for 1.09 deaths per 1000 patients.^2^ The economic burden of medication-related harm is estimated at $30.1 billion and 79 billion euros in United States and Europe respectively.^3^ Medication-related harm, also termed as adverse drug events (ADEs) include preventable or non-preventable harm caused by interventions related to medication use.^4^ Preventable medication error can occur at any step from the physician prescribing medications to the patient receiving the medication. In turn, ADEs are under active surveillance during healthcare delivery to patients with health systems or pharmacovigilance activities.

Advances in artificial intelligence (AI), digitization of health records, and accessibility to electronic patient records have been shown to reduce the occurrence, duration, and severity of ADEs.^5–7^ An AI-powered system has been reported to reduce inpatient prescribing error by up to 20%.^8^ However, traditional predictive models are still limited by the lack of in-depth clinical reasoning, poor interoperability in electronic health record systems (EHRs), difficulty in detecting rare events or interactions, and paucity of models that leverage unstructured data. Based on existing system, overlooked ADEs would lead to significant healthcare complications while trivial or clinical insignificant effects are over emphasized leading to healthcare administrative burden. Thus, with significant promises to address such unbalanced issue, generative AI (GenAI) and large language models (LLMs) may enable novel approaches previously unfeasible with conventional methods. For instance, preliminary studies have explored the potential of ChatGPT to recognize adverse drug reactions^9^, pharmacovigilance signal detection^10^, and automated medication chart review.^11,12^

This systematic review summarizes the breadth and depth of existing literature on how generative AI have been utilized to reduce ADEs and highlights areas for future investigation.

## Methods

### Search Strategy and Selection Criteria

This systematic review was conducted according to PRISMA guidelines.^16^ We searched PubMed, Web of Science, Embase, and Scopus to identify studies published between 1^st^ January 2012 to 18^th^ February 2024, related to application of generative AI in reducing medication related harm. Details of the search terms are provided in [Supplement III].

Studies were included if they were published in English, described the development or application of generative AI in mitigating potential medication related harms in the care delivery process (Figure 1), and were peer-reviewed original research, review and viewpoints, structured reviews of the literature reported in accordance with PRISMA guidelines, conference abstracts, case reports. We excluded studies that utilized solely predictive modeling approaches or investigated ADEs related to dietary supplements or use of medication not prescribed for the individual.

**Figure 1.**
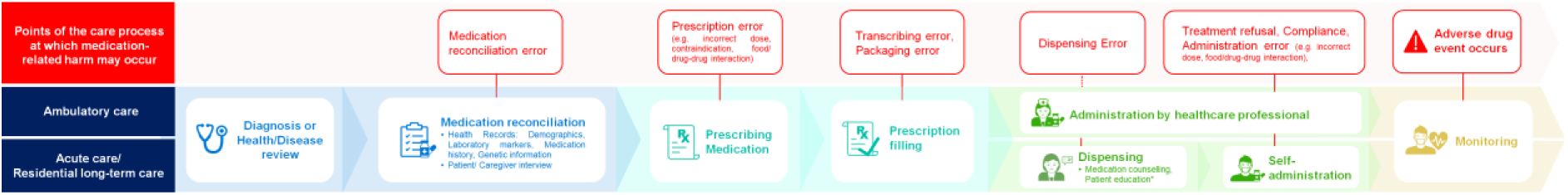
Archetypical care delivery process and potential points of error.

Based on these criteria, abstracts were screened for eligibility by two independent reviewers using a standardized tool. If no exclusion criteria were apparent in the abstract, it was included for manuscript review. Full-text manuscripts were conducted by two independent reviewers. Studies that did not meet the selection criteria were excluded at this stage. In cases of discrepancy between reviewers, eligibility was determined by a third reviewer.

### Data Analysis

We used a standardized form to extract pertinent information, including study characteristics, model details, application setting, outcome measures, findings, and reported challenges and limitations [Supplement IV]. We did not perform a critical appraisal of study quality as our primary objective is to characterise the scope of research in this field, identify research trends and gaps. The wide range of study designs and outcomes also precluded the application of a uniform quality assessment criterion.

## Results

The search yielded 2203 articles from all databases, with 1734 remaining after removing duplicates. After applying inclusion and exclusion criteria, 14 articles were eligible for this review.

**Figure 1a:**
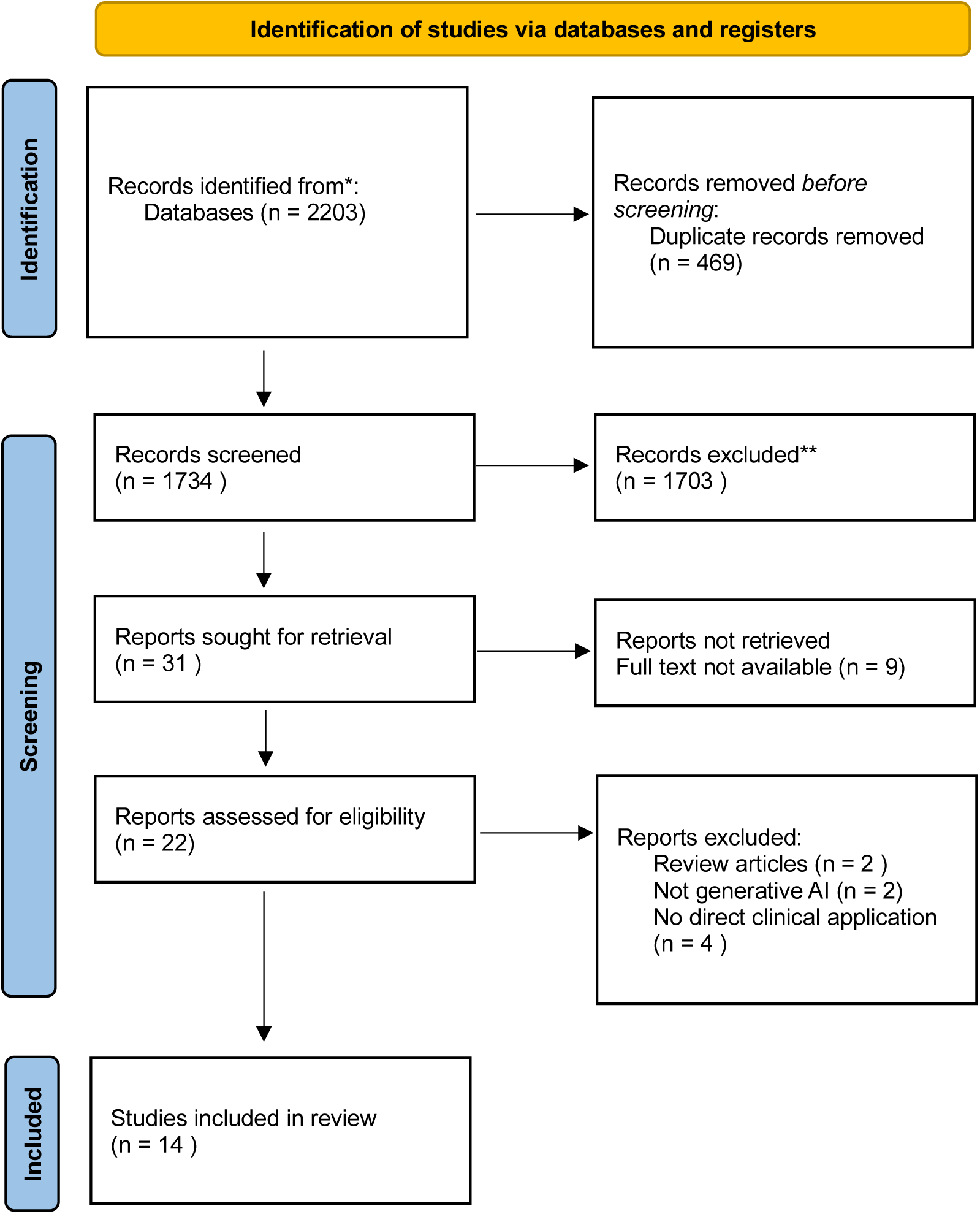
PRISMA flow diagram^17^

### Study Characteristics

Studies evaluated the performance of GenAI models for various applications, as summarized in Table 2. Four studies focused on the identification, classification, or prediction of drug-drug interactions (DDI).^18–21^ Three studies assessed the performance and utility of GenAI as decision support tools in benzodiazepine deprescribing^22^, aid dosing calculation of crushed tablets^23^ and provision of drug information^24^. Majority of studies focused on the application of GenAI in adverse event monitoring from specific drug classes and enhancing pharmacovigilance processes. Study designs were predominantly observational and cross-sectional. None of the studies tested models prospectively in their respective settings of application. The proposed applications of GenAI were broadly distributed across clinical (community, inpatient care) and public health settings.

**Table 1.** Key concepts and definitions.

| Concepts | Definitions |
| --- | --- |
| **Medication related harm** | The harm caused by medication if taken incorrectly, monitored insufficiently or as the result of an error, accident or communication problem.<sup>13</sup> |
| **Adverse drug event** | Any injury resulting from medical interventions related to a drug. This includes both adverse drug reactions in which no error occurred and complications resulting from medication errors.<sup>14</sup> |
| **Pharmacovigilance** | The science and activities relating to the detection, assessment, understanding and prevention of adverse effects or any other drug-related problem.<sup>15</sup> |
| **Generative artificial intelligence** | Generative AI: Deep-learning based AI techniques that can be used to create or produce various types of new contents, including text, images, audio, and videos, based on patterns learnt from training data. This includes foundation models and large language models. |

**Table 2:**
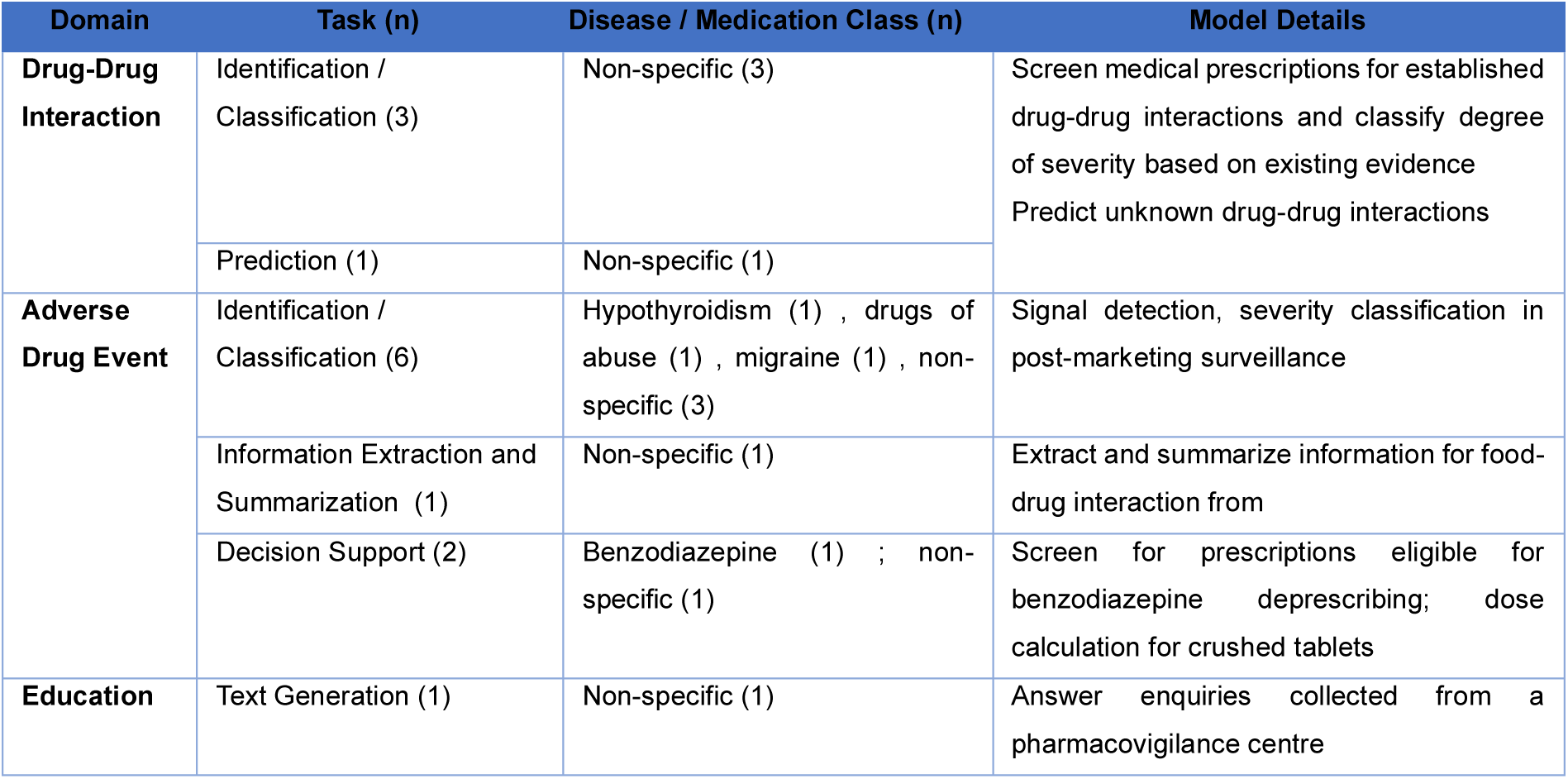
Summary of studies included in review.

| Domain | Task (n) | Disease / Medication Class (n) | Model Details |
| --- | --- | --- | --- |
| **Drug-Drug Interaction** | Identification / Classification (3) | Non-specific (3) | Screen medical prescriptions for established drug-drug interactions and classify degree of severity based on existing evidence<br>Predict unknown drug-drug interactions |
|  | Prediction (1) | Non-specific (1) |  |
| **Adverse Drug Event** | Identification / Classification (6) | Hypothyroidism (1) , drugs of abuse (1) , migraine (1) , non-specific (3) | Signal detection, severity classification in post-marketing surveillance |
|  | Information Extraction and Summarization (1) | Non-specific (1) | Extract and summarize information for food-drug interaction from |
|  | Decision Support (2) | Benzodiazepine (1) ; non-specific (1) | Screen for prescriptions eligible for benzodiazepine deprescribing; dose calculation for crushed tablets |
| **Education** | Text Generation (1) | Non-specific (1) | Answer enquiries collected from a pharmacovigilance centre |

**Table 3:** Types of generative models and data characteristics.

| Generative Model | Type(s) of Data Input | Training Datasets (if used) | Performance Metrics |
| --- | --- | --- | --- |
| **Large Language Models**<br>(ChatGPT-3.5, ChatGPT-4, Bing AI, Google Bard) | Structured, unstructured and combination of both structured and unstructured data | Studies did not use any training datasets (zero-shot inference) | Accuracy, Sensitivity, Specificity, Negative Predictive Value, Positive Predictive Value, readability, cohen's kappa, F-1 statistic, Precision, Recall |
| **Generative Adversarial Network** | Unstructured and combination of both structured and unstructured | Spontaneous ADR reports from pharmacovigilance centres, DrugBank, WebMD | Accuracy, AUPR, AUROC, Macro-F1, Precision, Recall, ROUGE, Likert Scale for qualitative human expert evaluation |

### Dataset Characteristics

Pharmacovigilance tasks often used public health databases, including FDA Adverse Event Reporting System (FAERS) Public Dashboard^25^, Health Canada ADR reporting dashboard^26^, China Food and Drug Adminstration^27^. Various datasets were used for named entity recognition tasks in ADE detection, such as user-generated content from web platforms (Medicitalia^28^, WebMD^29^) and open or restricted access datasets (CoNLL2003^30^, BioCreative V CDR^31^, n2c2^32^). Models built for DDI prediction utilized closed-source or in-house datasets such as DrugBank^24,33^. Testing datasets are often accrued from prior published studies, such as drug-drug interaction and deprescribing case scenarios^22,34^ with one study using an in-house retrospective cohort of medication prescriptions.^19^

### Model Types

Proprietary LLMs featured frequently in the reviewed studies, including various versions of ChatGPT and Google Bard. Studies used simple prompts or iterative prompting to generate responses on pretrained LLMs. None of the studies reported the use of additional techniques to enhance model performance, such as retrieval augmented generation or fine-tuning.

Custom-developed models adopt iterations of generative adversarial networks (GAN) and variational autoencoders (VAE) in the studies reviewed. The GTCACS^35^ approach was a three-step approach to better identify discussion topics from social media texts. GAN achieved dimensionality reduction, keyword clustering and summarization. DeepSAVE^36^, a deep learning framework used an enriched VAE for dimensionality reduction through parsimonious modelling of events captured on social media platform. In one study, BART (Bidirectional and Auto-Regressive Transformers) fine-tuned with a small amount of ADR-specific named entities (few-shot learning) was adapted to allow automated identification of diverse ADEs using small volumes of annotated data.^37^ GAN was adapted in DGANDDI^21^ into a graph attention network that encode drug attributes. DGANDDI was capable of binary and multi-class prediction tasks for drug-drug interactions using an enhanced and augmented multi-dimensional dataset generated by GAN. In a similar fashion, GAN was used to generate artificial features to augment data distribution in an imbalanced spontaneous reporting dataset.^27^

### Model Performance

For tasks that assist clinical decision making, reference standards include information from knowledge databases e.g. Lexicomp^®^ and expert opinion (healthcare providers or pharmacologists). Most commonly reported metrics include accuracy, sensitivity, specificity, F-1 statistic, precision, recall, AUC and AUPRC. Bespoke metrics include qualitative assessment of model responses by human experts, graded on Likert scales for quality, completeness or satisfaction. One study used ChatGPT-4 for qualitative evaluation of model performance, performed in parallel with human expert evaluation.^38^

#### Drug-Drug Interaction Classification and Prediction^18–21^

In prediction of potential drug interaction pairs, GAN-based models achieved high accuracy rate. GANs are generative models that learn from the distribution of data or images to create large, realistic synthetic data.^39^ DGANDDI outperformed baseline methods in both binary and multi-class DDI prediction tasks. In binary prediction, it achieved an accuracy of 96.10%, AUPR of 99.27%, and AUROC of 99.26%. In the multi-class prediction task, DGANDDI attained an accuracy of 95.89%, AUPR of 97.29%, and AUROC of 99.97%.

Proprietary LLMs were used to classify DDIs. One study compared the performance of different LLMs including Microsoft Bing AI, ChatGPT-3.5 and ChatGPT-4 and Google Bard.^18^ When Micromedex was used as the reference standard, accuracy of LLMs ranged between 0.469 to 0.788, with Microsoft Bing AI demonstrating the best performance. Sensitivity was comparable across all LLMs, but specificity was significantly lower for ChatGPT-3.5 and ChatGPT-4. In another study, the Google Bard was used to screen prescriptions for drug-drug interactions, demonstrated low degree of agreement with predictions from Lexicomp.^19^ There was a nil to slight agreement between interaction risk rating (κ=0.01), severity rating (κ=0.02), and reliability (κ =-0.02). Conversely, ChatGPT (version not reported) was found to be highly accurate in identifying drug-drug interactions in 39 out of 40 DDI pairs tested. When prompted to explain its answer, ChatGPT produced responses that was highly readable.

#### Decision Suppor^t22–24^

In decision support applications, a study leveraging GPT-4 for benzodiazepine deprescribing reported high degree of overall agreement between LLM and human expert in identifying cases eligible for deprescribing.^22^ Agreement on four different deprescribing criteria was varied, ranging 74.7% to 91.3% (lack of indication: κ = .352, P < .001; prolonged use: κ = .088, P = .280; safety concerns: κ = .123, P = .006; incorrect dosage: κ = .264, P = .001). Qualitative analysis of GPT-4 responses found that up to 22% were ambiguous, generic and contained inconsistencies. Another study introduced a web-based calculator developed to guide dosing calculation, particularly in paediatric care where such errors are prevalent.^23^ The authors used ChatGPT (version not reported) and Visual Studio to write the underlying HTML code for dose division calculations and webpage interface creation. The webpage’s reliability and feasibility were then assessed using retrospective data and validated questionnaires, scoring 88.38 on the System Usability Scale. Accuracy and reproducibility of the calculator was not evaluated.

In the provision of pharmacovigilance related enquiries, ChatGPT-4 responses was compared against responses by pharmacovigilance specialists. The median score (IQR) of the ChatGPT’s responses on a 10-point Likert scale was 4.8 (3–7.3), with a specific focus on drug causality scoring lower at 3.7 (3– 6.3), and information on medication and proper use scoring slightly better at 5 (3.2–8.3). The authors conclude that chatbot’s responses were generally not acceptable, especially in terms of precision and clinical relevance.

#### Pharmacovigilance^27,35–37,40^

For signal detection and ADR classification, studies used generative AI for training data augmentation and dimension reduction. These models e.g. GTCACS outperformed non GenAI methods on internal validity measures. The DeepSAVE model, which uses a VAE approach, was tested on a dataset comprising of 104 million user search queries and 800 events. DeepSAVE outperformed existing methods (e.g. disproportionality analysis, DA atop Event Mention Classifier) with the highest F-measure across all validation datasets. A GAN-based classification model developed to automatically evaluate risk categories of drugs during post-marking surveillance demonstrated highest accuracy of 97.9% when compared against existing models.

In one study, authors demonstrated few-shot learning with LightNER and BART, the ADR recognition performance in low-resource datasets significantly improved. For instance, the LightNER model fine-tuned using the N2C2 dataset, achieved an F1-score of 61.42%, indicating the model’s effective transfer of task knowledge from rich-resource to low-resource settings. GPT-3 was used in another study to generate a comprehensive lexicon of drug abuse synonyms from social media sources. Coupled with automated API queries and simple automated filters (e.g. google filters), the proposed method yielded precision of 0.859 and 0.770, recall of 0.431 and 0.395 for alprazolam and fentanyl respectively.

LLM was harnessed to improve efficiency of pharmacovigilance process in one study, where authors used iterative prompting of GPT-4 to review and summarize food effects on drugs from drug review documents. Final draft summaries generated by GPT-4 were rated by FDA professionals, with 85% rated as factually consistent with reference summaries. This showcases GPT-4’s potential to aid in faster and more reliable drug assessment processes.

## Discussion

As healthcare systems increasingly prioritize patient safety, the integration of AI has the potential to enhance the detection and prevention of ADEs, and, by extension, reduce the substantial economic cost. Our scoping review revealed three key applications of GenAI in the literature to date: identification and prediction of drug-drug interactions, provision of decision support in medication management, and automation of pharmacovigilance activities.

### Effectiveness of Generative AI in Enhancing Safety

Drug-drug interactions make up nearly 3% of all hospital admissions and account for up to 5% of all inpatient medication errors.^41^ Harmful DDIs are often only reported from post-marketing surveillance activities, rather than at the clinical trial stage.^42^ Our review included studies that predict potential DDIs pre-clinically. Performance of models augmented by GAN outperforms those trained using traditionally augmented data using a fraction of the original training dataset.^43,44^ GAN can be a useful tool in enhancing prediction accuracy where data is limited. On the other hand, LLMs demonstrated variable performance in screening DDIs from prescriptions. Studies frequently used simple prompting strategies to elucidate response from LLMs, with no additional techniques used to provide contextual knowledge or reduce incorrect responses (or “hallucinations”). Methods such as retrieval augmented generation (RAG) or fine-tuning may allow LLMs to tailor responses to specified tasks through provision of contextual knowledge (e.g. drug-drug interaction database).^45,46^ The advantage of such techniques have been shown in other clinical tasks, including differential diagnosis^47^, evidenced-based decision support^46,48^ and patient chart review^49^. These techniques however, may rely on well-curated, clinically adjudicated drug-drug interaction datasets that are not often freely available.

As decision support tools, studies adopting LLMs are mainly exploratory in nature. We found a wide range of tasks and purposes (e.g. prescription review, dosage calculation, and answering medication enquiry). These broad applications are enabled by generalist properties of large language models.^50^ LLMs demonstrate capacity to perform tasks with little to no task-specific training, also known as “zero-shot” or “few-shots” learning. In the context of reducing medication harm, LLMs may simulate clinical reasoning and inferential skills across diverse medical disciplines, drug classes and user settings without the need for explicit training. For instance, an LLM trained to screen prescriptions for inappropriate benzodiazepine use may be adapted easily to screen for inappropriate drug use in elderly patients. In addition, LLMs are well poised as medical chatbots, given their text generation capabilities demonstrating high degree of fluency, empathy, and personalization, even outperforming clinicians.^51^ These explorations, however, highlight existing challenges to clinical adoption of LLMs. While studies to date are in research phase and no exploration in terms of auto-piloting or co-piloting as modes of clinical integration. Accuracy, reliability and consistency of LLM responses using general purpose LLMs such as ChatGPT precludes its autonomous use in clinical settings.

A promising area of LLM application in enhancing efficiency and impact is in ADE monitoring and pharmacovigilance, where GenAI tools may enhance timeliness and accuracy of ADR detection from specific medication classes. Our review has shown that GenAI enhanced accuracy of signal detection, disease and drug entity recognition over conventional natural language processing tools. LLMs are able to handle a wide breadth of data sources (i.e. electronic health records, online databases, and social media platform), facilitating the detection of rare events and offering a generalist capability that is essential for continuous learning and adaptation.^52^ Automation of specific tasks in pharmacovigilance that is traditionally resource-intensive is a potential avenue for productivity gain with the use of GenAI models.^53^

### Clinical Implications

In our review, we are unable to provide conclusive evidence that GenAI will reduce medication-related harms when applied in clinical settings. Along the continuum of medication use process, GenAI models were actually only adopted for highly selective domains and tasks. While a comprehensive review or metrics of medication-related tasks across multiple domains or tasks across different GenAI models have yet to be concluded. In other words, current generative AI in research are still applied as narrow based AI focused on specific task, yet to be explored or achieve their “generative” potential for medication safety. Non-generative AI models was evaluated in another review, where 78 articles described the application of AI in reducing ADEs.^54^ A variety of AI techniques were described including neural networks and tree-based algorithms in predicting potential ADEs and enhancing early detection.

Utilizing diverse data sources like genetic information and electronic health records, these AI models aimed to inform clinical decisions on safe prescribing and medication management.

Instead of applying generative AI models in tasks that mandate deterministic outputs, we propose that LLMs can be adopted in ways to reduce cognitive workload for healthcare professionals. Healthcare professionals work with high volumes of multi-modal patient data and are required to pay attention to details, synthesize information and make clinical decisions in real time. High cognitive load pose risk for burnout and medical errors.^55^ For example, LLMs can be used to analyse and reduce alert burden in electronic medical records, in medication incident analysis, and summarization in a similar fashion to discharge notes generation.^12,56^ In a study published after we completed literature search, LLMs were used in a co-pilot system to extract key named entities of online submitted prescriptions and assembly into coherent instructions.^57^ This system was shown to reduce near-miss events and improved the efficiency of pharmacy operations in a large-scale online pharmacy. Finally, LLMs can be leveraged upon as a tool in patient education and engagement thereby enhancing patient access to critical medication related information.^58^

A critical evaluation of studies included in our review revealed a lack of adherence to reporting guidelines for AI studies. We did not perform a quality review of the studies in view of the scoping nature of this review and diverse hypotheses of studies included. Checklists and reporting guidelines such as the MI-CLAIM for transparent model reporting^59^, TRIPOD+AI checklist for comprehensive reporting of predictive models^60^ and DECIDE-AI checklist for early stage clinical evaluation of AI-based decision support tools^61^ should be adopted in future studies. However, there is still lack of validated reporting tools for LLM-based AI model, though initial efforts have been made to create LLM-specific frameworks.^62^ Evaluation or discussions on model fairness, bias and other ethical considerations such as data privacy were also found to be lacking in the included studies.

### Limitations

Our study has several limitations. The heterogeneity across studies regarding application, GenAI tools used and setting prevented a formal assessment of predictive validity for different AI models. Diversity of training and testing datasets used precludes generalizability of findings across different demographic groups. Patient outcomes were not reported in all studies, limiting any conclusions about the role of GenAI and its impact on patient outcomes. We limited our review to only peer reviewed articles. We acknowledge that the field of generative AI and LLM is rapidly evolving and a large number of studies may still be in the preprint stage or archived.

### Blueprint for Future Studies

Future research should focus on developing and benchmarking generative AI models against established healthcare standards to further validate their performance and cost-effectiveness to ensure their safe integration into clinical practice. There is a need to develop expert curated, high quality training datasets with diverse representation from different geographical, ethnic and social groups. Such datasets, when shared, can facilitate and accelerate training of GenAI models adapted for different applications in the medication use process. For example, an expert annotated dataset of incident reports can be used to fine-tune an LLM-based model to predict risk for medication incidents.^63^ Other areas of high interest include the use of LLMs for real-time monitoring of drug safety and the exploration of GAN the synthetic generation of training data, which can help overcome the limitations posed by rare ADE occurrences.

## Conclusion

GenAI and LLMs demonstrate potential in enhancing medication safety and reducing medication-related harm. Published studies reveal potential areas for successful future implementation. However, the current areas that have been addressed are targeted at only some of the key safety issues of medication safety today. Moreover, research rigor and comprehensive ethical evaluation is lacking in the studies to date. Future studies should address gaps in lack of high-quality datasets specific for medication safety tasks. Continuous update of this review is warranted given the burgeoning nature of this field.

## Contributors

NL, JCLO and CM conceptualized and designed the systematic review. JCLO, CM, NN, KB, NYTT, LJ and QX performed article screening and data extraction. RR, DWB, DSWT contributed to the first draft of the report with input from NL. All authors had full access to all the data in the study and had final responsibility for the decision to submit for publication.

## Data Availability

All data produced in the present study are available upon reasonable request to the authors

---

**Supplement I.**
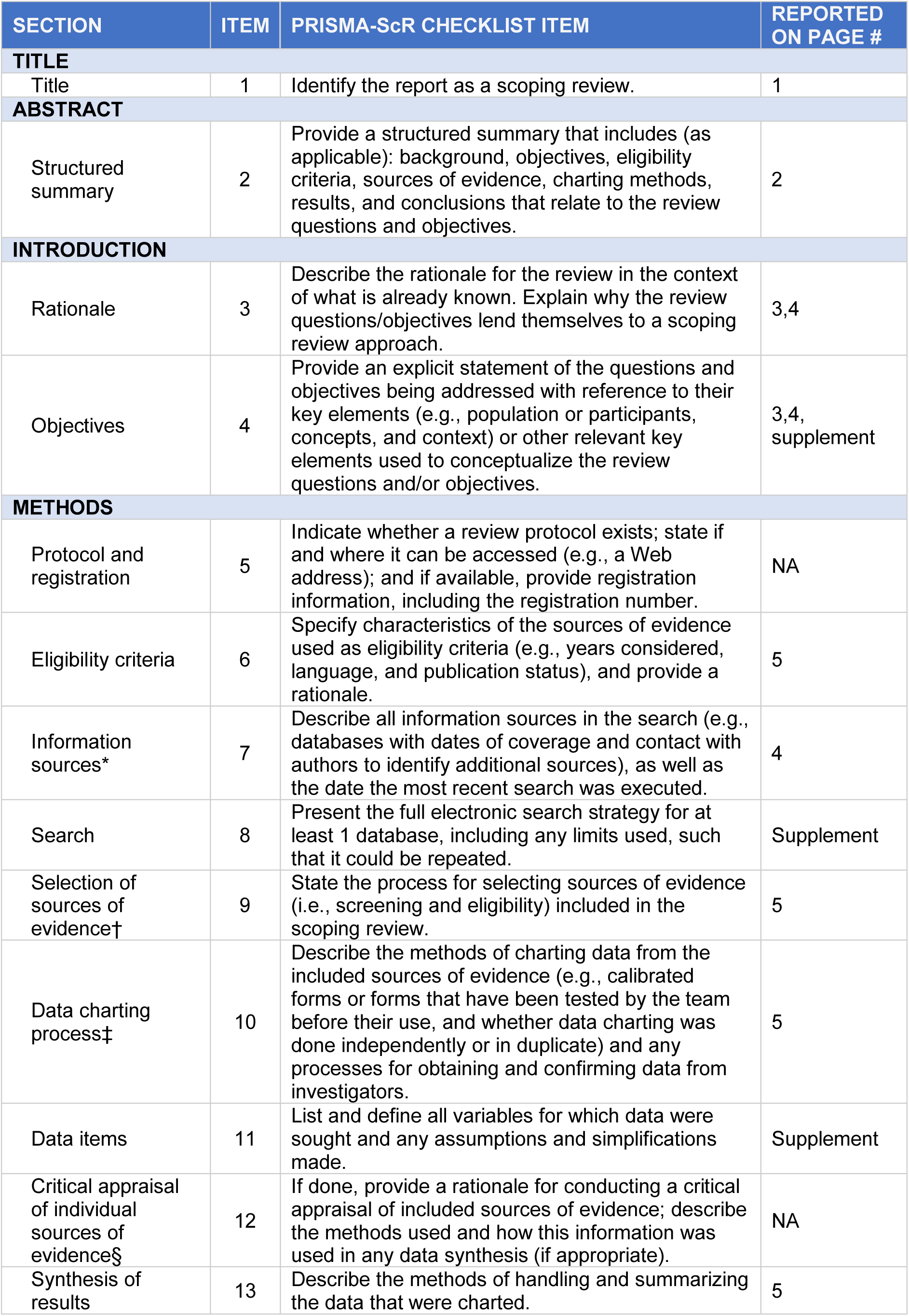
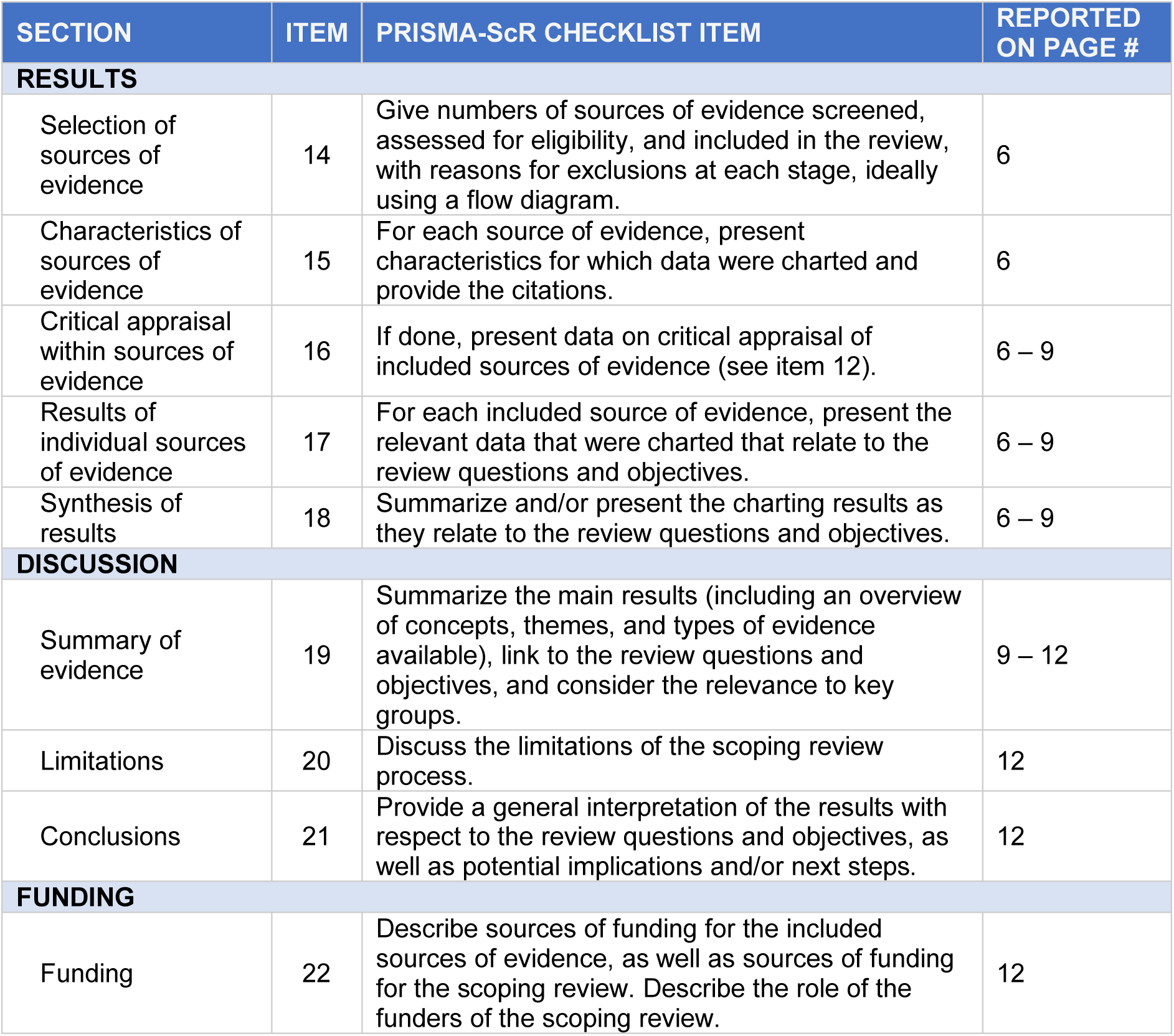
Preferred Reporting Items for Systematic reviews and Meta-Analyses extension for Scoping Reviews (PRISMA-ScR) Checklist.

| SECTION | ITEM | PRISMA-ScR CHECKLIST ITEM | REPORTED ON PAGE # |
| --- | --- | --- | --- |
| **TITLE** |  |  |  |
| Title | 1 | Identify the report as a scoping review. | 1 |
| **ABSTRACT** |  |  |  |
| Structured summary | 2 | Provide a structured summary that includes (as applicable): background, objectives, eligibility criteria, sources of evidence, charting methods, results, and conclusions that relate to the review questions and objectives. | 2 |
| **INTRODUCTION** |  |  |  |
| Rationale | 3 | Describe the rationale for the review in the context of what is already known. Explain why the review questions/objectives lend themselves to a scoping review approach. | 3,4 |
| Objectives | 4 | Provide an explicit statement of the questions and objectives being addressed with reference to their key elements (e.g., population or participants, concepts, and context) or other relevant key elements used to conceptualize the review questions and/or objectives. | 3,4, supplement |
| **METHODS** |  |  |  |
| Protocol and registration | 5 | Indicate whether a review protocol exists; state if and where it can be accessed (e.g., a Web address); and if available, provide registration information, including the registration number. | NA |
| Eligibility criteria | 6 | Specify characteristics of the sources of evidence used as eligibility criteria (e.g., years considered, language, and publication status), and provide a rationale. | 5 |
| Information sources* | 7 | Describe all information sources in the search (e.g., databases with dates of coverage and contact with authors to identify additional sources), as well as the date the most recent search was executed. | 4 |
| Search | 8 | Present the full electronic search strategy for at least 1 database, including any limits used, such that it could be repeated. | Supplement |
| Selection of sources of evidence† | 9 | State the process for selecting sources of evidence (i.e., screening and eligibility) included in the scoping review. | 5 |
| Data charting process‡ | 10 | Describe the methods of charting data from the included sources of evidence (e.g., calibrated forms or forms that have been tested by the team before their use, and whether data charting was done independently or in duplicate) and any processes for obtaining and confirming data from investigators. | 5 |
| Data items | 11 | List and define all variables for which data were sought and any assumptions and simplifications made. | Supplement |
| Critical appraisal of individual sources of evidence§ | 12 | If done, provide a rationale for conducting a critical appraisal of included sources of evidence; describe the methods used and how this information was used in any data synthesis (if appropriate). | NA |
| Synthesis of results | 13 | Describe the methods of handling and summarizing the data that were charted. | 5 |
| **RESULTS** |  |  |  |
| Selection of sources of evidence | 14 | Give numbers of sources of evidence screened, assessed for eligibility, and included in the review, with reasons for exclusions at each stage, ideally using a flow diagram. | 6 |
| Characteristics of sources of evidence | 15 | For each source of evidence, present characteristics for which data were charted and provide the citations. | 6 |
| Critical appraisal within sources of evidence | 16 | If done, present data on critical appraisal of included sources of evidence (see item 12). | 6 – 9 |
| Results of individual sources of evidence | 17 | For each included source of evidence, present the relevant data that were charted that relate to the review questions and objectives. | 6 – 9 |
| Synthesis of results | 18 | Summarize and/or present the charting results as they relate to the review questions and objectives. | 6 – 9 |
| **DISCUSSION** |  |  |  |
| Summary of evidence | 19 | Summarize the main results (including an overview of concepts, themes, and types of evidence available), link to the review questions and objectives, and consider the relevance to key groups. | 9 – 12 |
| Limitations | 20 | Discuss the limitations of the scoping review process. | 12 |
| Conclusions | 21 | Provide a general interpretation of the results with respect to the review questions and objectives, as well as potential implications and/or next steps. | 12 |
| **FUNDING** |  |  |  |
| Funding | 22 | Describe sources of funding for the included sources of evidence, as well as sources of funding for the scoping review. Describe the role of the funders of the scoping review. | 12 |

**Supplement II.**
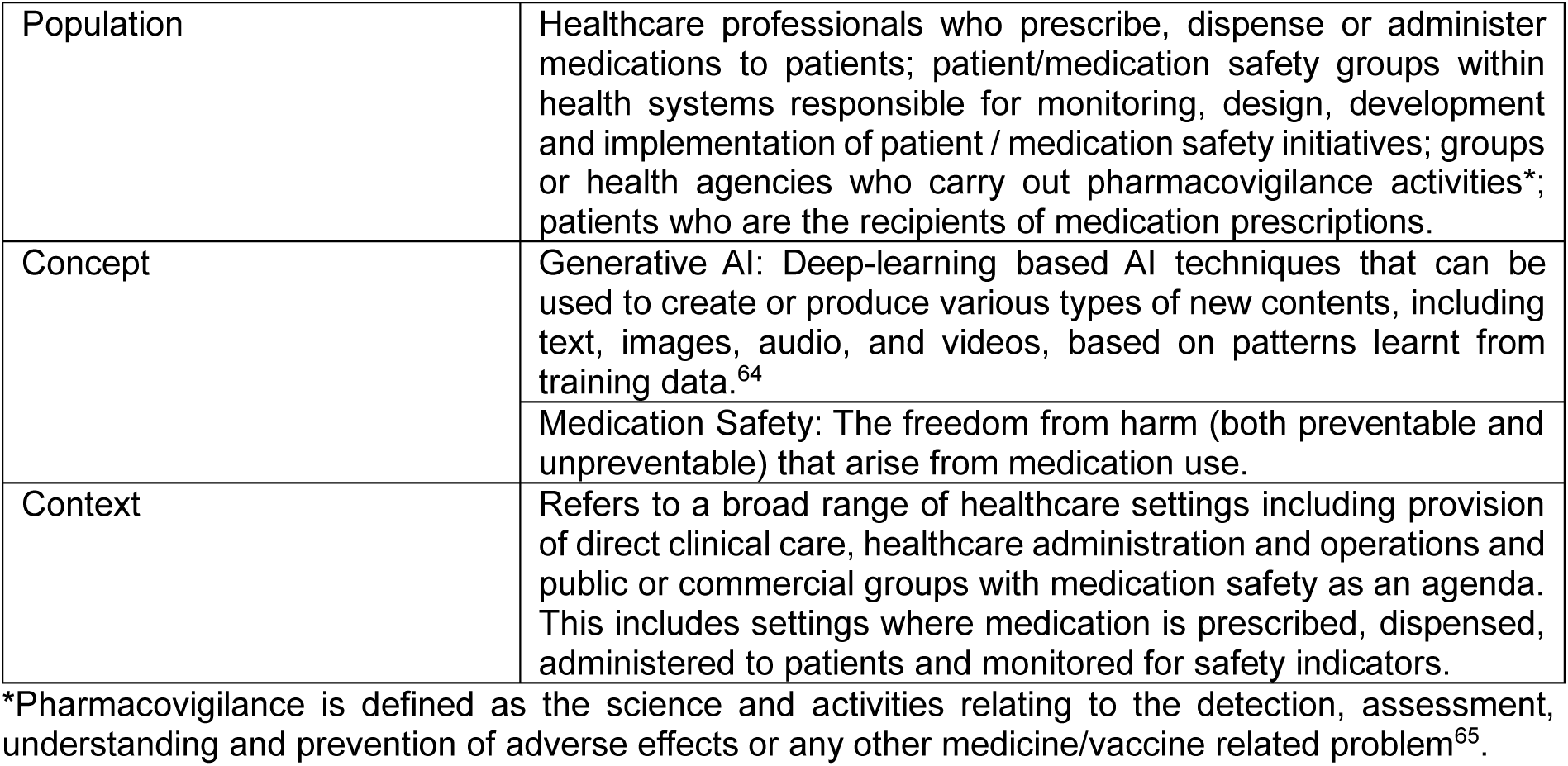
Population, Concept and Contexts (PCC) for the scoping review.

|  |  |
| --- | --- |
| Population | Healthcare professionals who prescribe, dispense or administer medications to patients; patient/medication safety groups within health systems responsible for monitoring, design, development and implementation of patient / medication safety initiatives; groups or health agencies who carry out pharmacovigilance activities*; patients who are the recipients of medication prescriptions. |
| Concept | Generative AI: Deep-learning based AI techniques that can be used to create or produce various types of new contents, including text, images, audio, and videos, based on patterns learnt from training data.<sup>64</sup> |
|  | Medication Safety: The freedom from harm (both preventable and unpreventable) that arise from medication use. |
| Context | Refers to a broad range of healthcare settings including provision of direct clinical care, healthcare administration and operations and public or commercial groups with medication safety as an agenda. This includes settings where medication is prescribed, dispensed, administered to patients and monitored for safety indicators. |

\*Pharmacovigilance is defined as the science and activities relating to the detection, assessment, understanding and prevention of adverse effects or any other medicine/vaccine related problem<sup>65</sup>.

**Supplement III.**
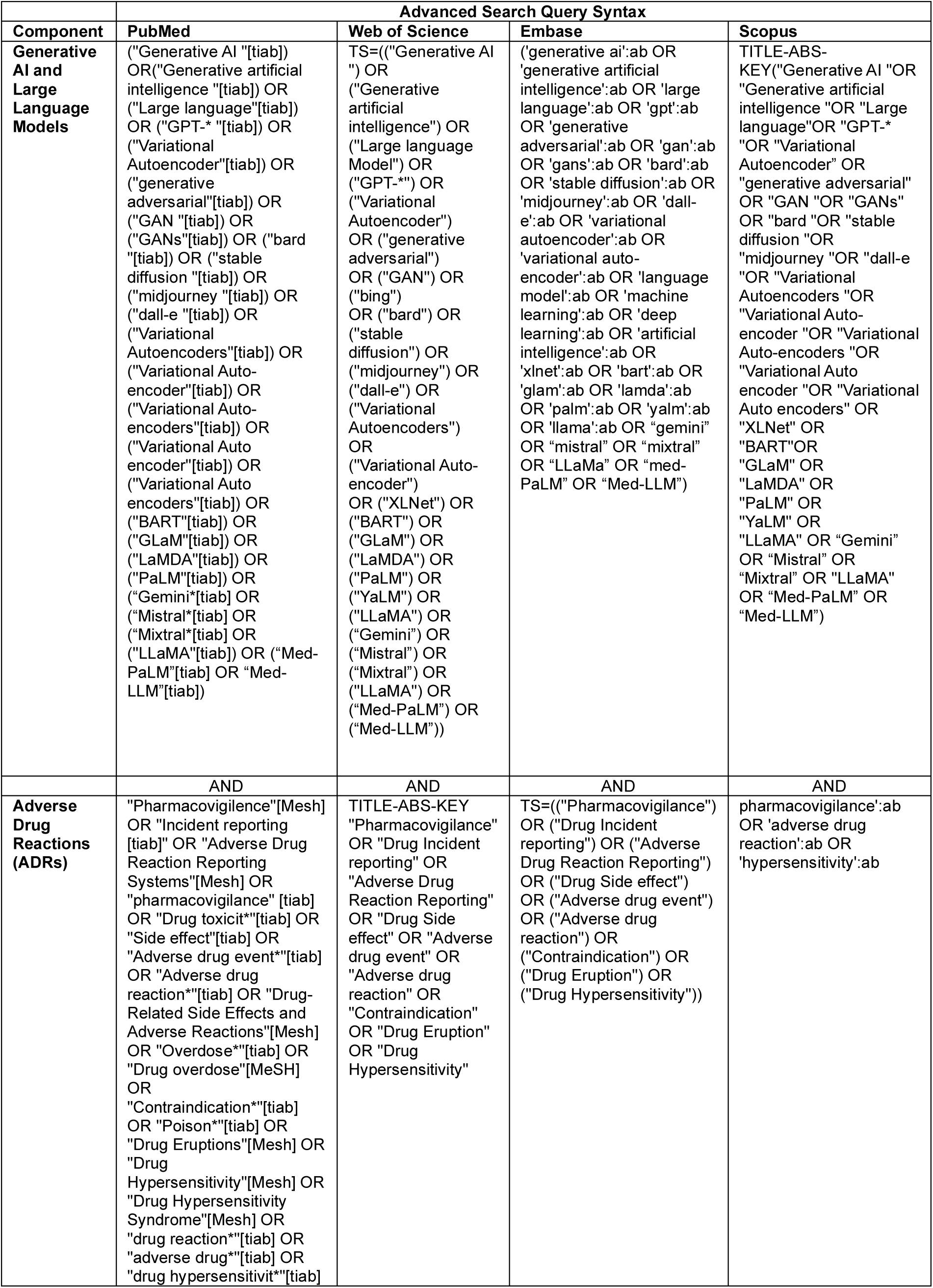
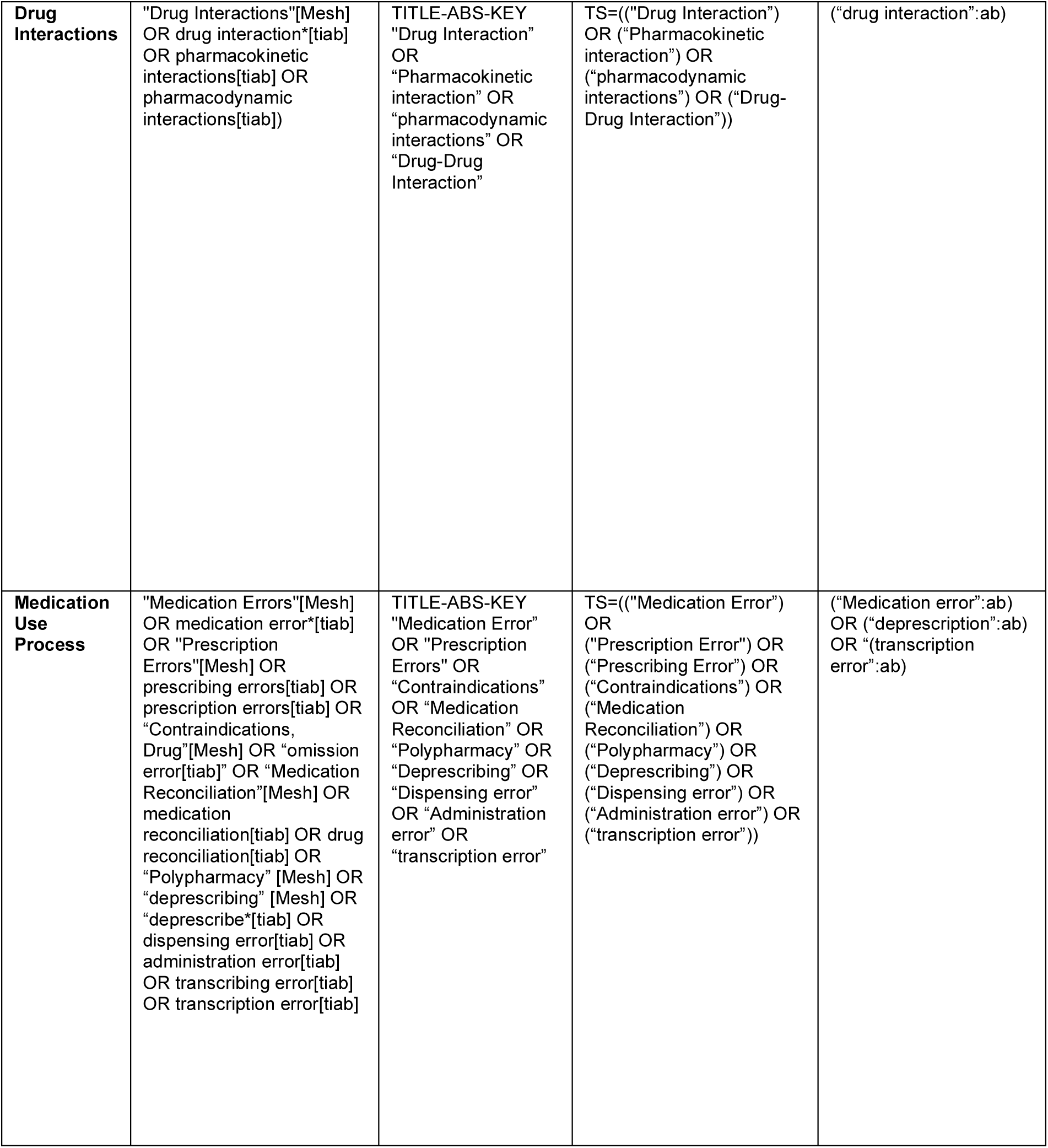
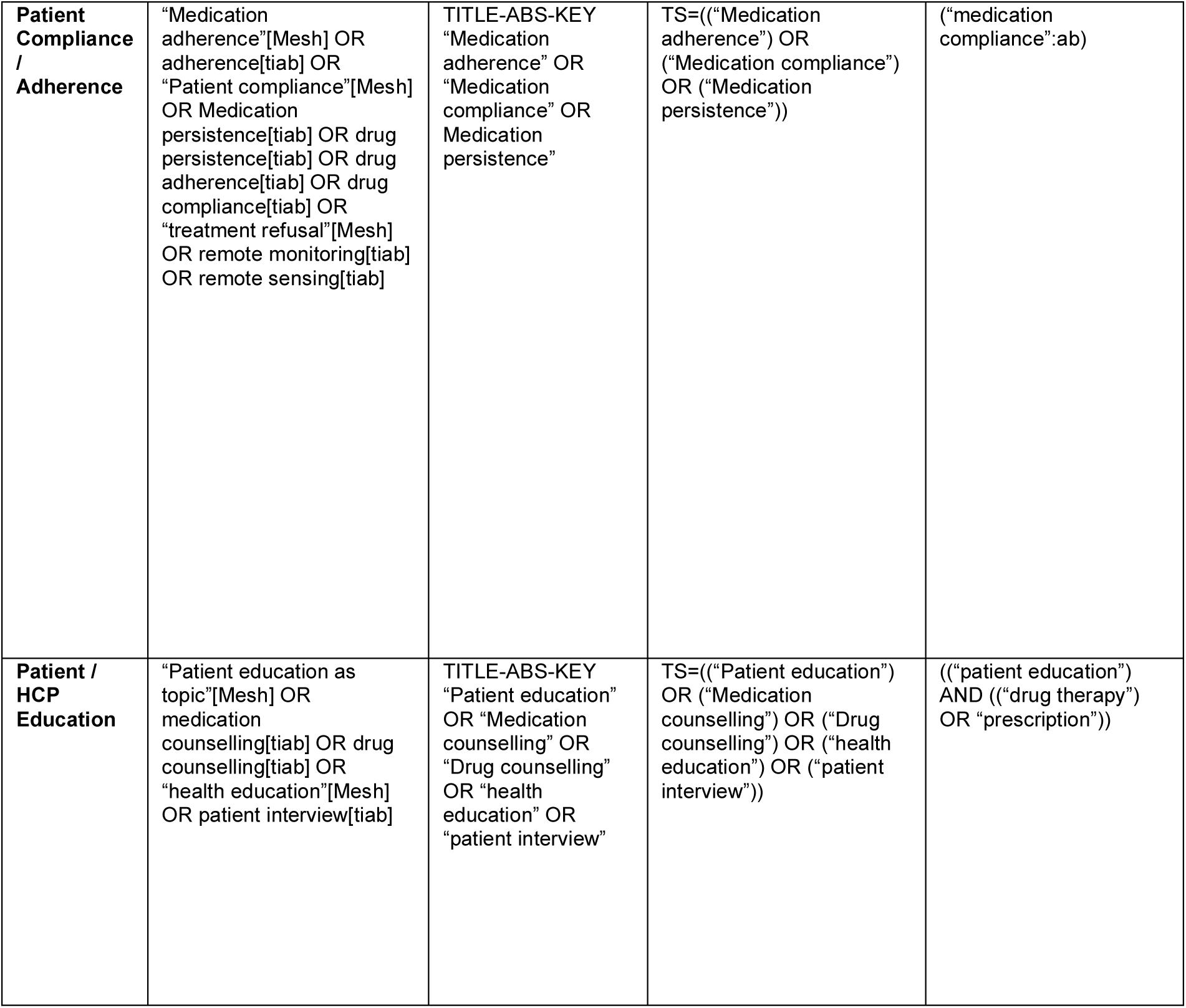
Search Strategy.

**Supplement IV.**
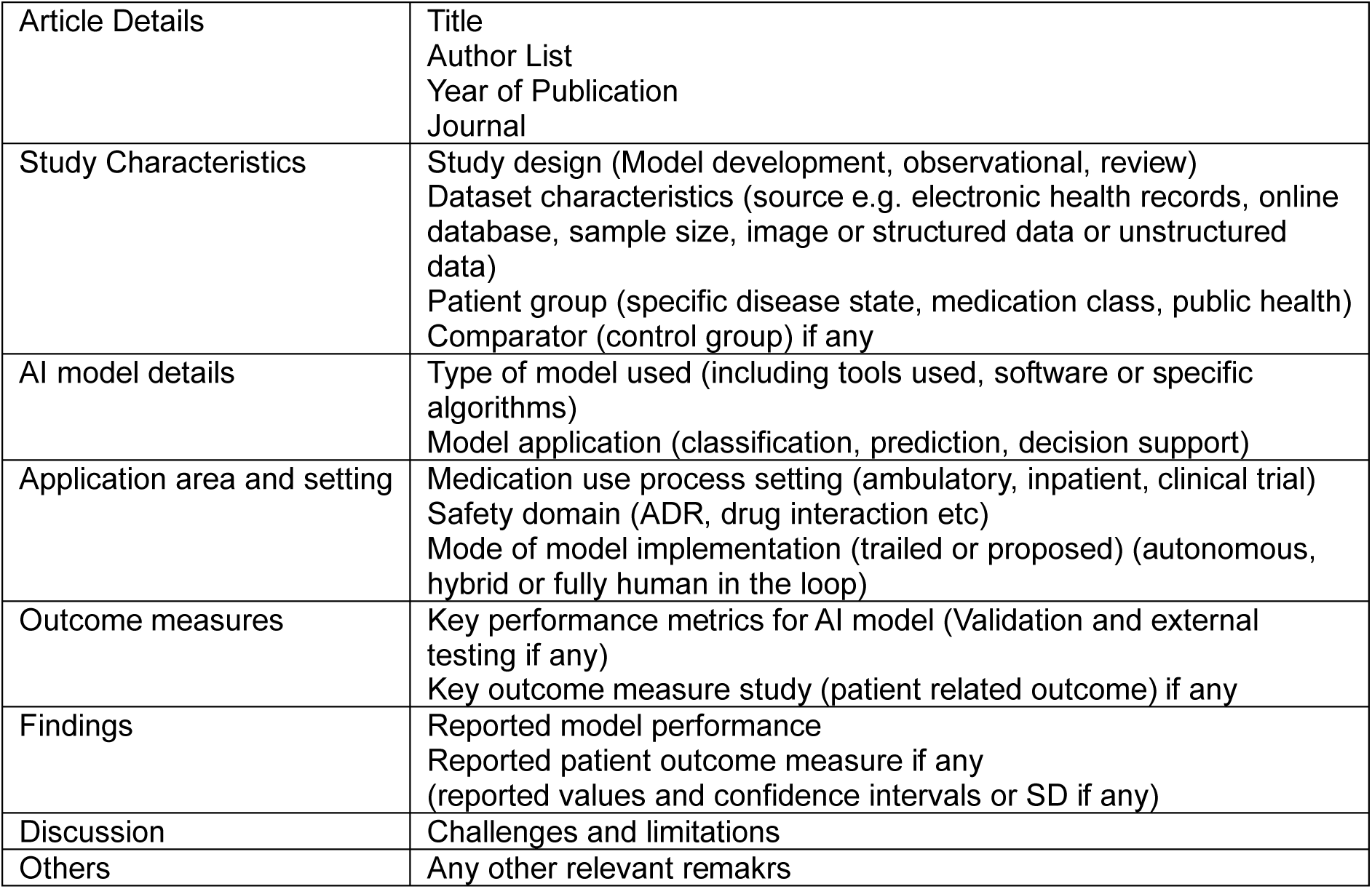
Data abstraction variables.

|  |  |
| --- | --- |
| Article Details | Title<br>Author List<br>Year of Publication<br>Journal |
| Study Characteristics | Study design (Model development, observational, review)<br>Dataset characteristics (source e.g. electronic health records, online database, sample size, image or structured data or unstructured data)<br>Patient group (specific disease state, medication class, public health)<br>Comparator (control group) if any |
| AI model details | Type of model used (including tools used, software or specific algorithms)<br>Model application (classification, prediction, decision support) |
| Application area and setting | Medication use process setting (ambulatory, inpatient, clinical trial)<br>Safety domain (ADR, drug interaction etc)<br>Mode of model implementation (trailed or proposed) (autonomous, hybrid or fully human in the loop) |
| Outcome measures | Key performance metrics for AI model (Validation and external testing if any)<br>Key outcome measure study (patient related outcome) if any |
| Findings | Reported model performance<br>Reported patient outcome measure if any<br>(reported values and confidence intervals or SD if any) |
| Discussion | Challenges and limitations |
| Others | Any other relevant remarks |

